# Development of an algorithm for the Charlson Comorbidity Index using the National Database Study in the Japan Kanto 7 Prefectures

**DOI:** 10.1101/2025.04.10.25325568

**Authors:** Airi Sekine, Kei Nakajima

## Abstract

The Charlson Comorbidity Index (CCI) contains 19 diseases, is weighted according to their potential influence on mortality, and has been widely used as a valid predictor for all-cause mortality. In this study, we aimed to develop an algorithm to define the CCI using information about diagnosed diseases and prescribed medication from the Japan National Database of Health Insurance Claims and Specific Health Checkups (NDB). The present study included 10,183,619 individuals who underwent health checkups from April 2018 to March 2019. We identified diagnosed diseases using Japan’s disease codes, which correspond to a more detailed disease classification than the International Classification of Diseases 10th edition codes, and Japan’s medication codes, which detail medication classifications.

The mean CCI scores of men, women, and all subjects were 0.67 ± 1.20, 0.68 ± 1.14, and 0.68 ± 1.17, respectively. For all patients, chronic pulmonary disease was the most prevalent condition out of the 19 Charlson comorbidities.

Although further validation studies are needed to assess the association between our CCI and actual mortality in NDB, this index can be useful for epidemiological studies using a real-world database to adjust for patient comorbid conditions.

## Introduction

The Charlson Comorbidity Index (CCI) was developed by Charlson et al. in 1987 and has been widely used as a comorbidity index to determine the mortality rate (at one and ten years) in patients with multiple comorbidities [1]. The CCI contains 19 diseases, including diabetes with diabetic complications, congestive heart failure, peripheral vascular disease, chronic pulmonary disease, mild and severe liver disease, hemiplegia, renal disease, leukemia, lymphoma, metastatic tumors, and acquired immunodeficiency syndrome (AIDS). These conditions are weighted according to their potential influence on mortality and are assigned values of 1, 2, 3, or 6. Charlson et al. showed that the accumulative mortality rate attributable to these comorbidities increased stepwise with increasing CCI scores. Additionally, the CCI has been used as a confounding factor in several outcome studies to adjust for potential severity in patients [2,3].

Several studies have developed algorithms to define the CCI using claim databases based on the International Classification of Diseases, 9th and 10th revisions (ICD-9 and 10) codes [4,5]. However, the original CCI was designed for medical chart data and included criteria that were not discernable from ICD-10 codes. For example, patients with diet-controlled diabetes were excluded from the CCI criteria [1].

The Japan National Database of Health Insurance Claims and Specific Health Checkups (NDB) was provided as secondary data by Japan’s Ministry of Health, Labour and Welfare (MHLW). Japanese claims data (all diagnosed diseases and prescribed medications) and annual health checkup information are stored in the NDB [6], making it a valuable administrative database for epidemiological studies in Japan. In this study, we aimed to develop an algorithm that defined the CCI using diagnosed diseases and prescribed medications registered in the NDB to help future epidemiological studies adjust for patient comorbid conditions.

## Materials and Methods

### Study design and participants

This study was part of the National Database Study from the Kanto 7 Prefectures (NDB-K7Ps Study conducted in Tokyo, Kanagawa, Saitama, Chiba, Ibaraki, Gunma, and Tochigi, Japan) and aimed to investigate clinical factors primarily associated with cardiometabolic disease. The NDB contains data on disease diagnoses, prescribed medications, and annual health checkup information [6]. Details of the study concept and design have been described elsewhere [7]. Our study protocol was accepted by the MHLW in December 2020, and we received digitally recorded anonymous data in July 2022 (No. 2320). This study was conducted according to the guidelines of the Declaration of Helsinki and approved by the Institutional Review Board of the Ethics Committee of Japan Women’s University (No. 513). Details of the data processing used to link health checkup data with datasets comprising diagnosed diseases and prescribed medications have been described elsewhere [8]. This study included 10,183,619 individuals living in the seven prefectures of Kanto (listed above) who underwent specific health checkups between April 2018 and March 2019. These checkups are mandatory for people aged 40–74 years in Japan [9].

### Disease diagnosis and prescribed medications

Japan’s disease codes are used for insurance claims [10] and correspond to a more detailed disease classification than ICD-10 codes [11,12]. We identified diagnosed diseases for the CCI using Japan’s disease codes that correspond to the ICD-10. Similarly, we identified prescribed medications using Japan’s medication codes for insurance claims, which corresponded to detailed medication classifications, including the dosage, form, method of administration, trade name (whether it was an original or generic product), and the pharmaceutical company that manufactures the drug. These disease and medication codes from insurance claims were rigorously checked by medical clerks in each hospital, clinic, and dispensing pharmacy. Patients were defined as those who were diagnosed or prescribed medication at least once from April 2018 to March 2019.

### Charlson Comorbidity Index

The CCI was calculated as the sum of the weighted index (1, 2, 3, or 6 points) of 19 comorbidities according to their potential influence on mortality for diabetes with diabetic complications, congestive heart failure, peripheral vascular disease, chronic pulmonary disease, mild and severe liver disease, hemiplegia, renal disease, leukemia, lymphoma, metastatic tumors, and AIDS [1]. Detailed definitions of the 19 comorbidities using Japan’s disease codes, based on the ICD-10 codes selected by Quan and Ono [5,13], and other criteria using medication codes are shown in ***Table 1***. We defined diabetes without chronic complications as patients diagnosed with at least one of the 30 disease codes for this condition and prescribed at least one of 487 oral hypoglycemic agents or 61 insulin preparations without intravenous injections (vial formulations). Diabetes with chronic complications included patients with diabetic retinopathy, neuropathy or nephropathy, or those hospitalized with diabetic ketoacidosis (DKA) or hyperosmolar hyperglycemic syndrome (HHS). Patients with diabetes who were prescribed at least one of the 23 medications for diabetic retinopathy were included in the diabetic retinopathy group. We defined DKA or HHS patients as those who were diagnosed with DKA, HHS, or diabetic coma. In addition, we defined hospitalized DKA or HHS patients as those who were prescribed insulin preparations for intravenous injection. Mild liver disease was defined as chronic hepatitis or cirrhosis without portal hypertension. Moderate liver disease included chronic hepatitis or cirrhosis with portal hypertension, while severe liver disease included variceal bleeding. Metastatic tumors were defined as end-stage cancer or unknown primary cancers. Several definitions of the CCI, such as hospitalization, detailed symptoms regarding electrocardiogram (ECG) changes, New York Heart Association (NYHA) functional classifications, and serum creatinine levels were unavailable in our database (similarly described in ***Table 1***). In this study, the CCI was not adjusted for age.

**Table 1.** Definitions of Charlson Comorbidity Index.

| Points | Charlson comorbidities | Corresponding ICD-10 code | Number of diagnoses*1 | Other criteria in our study | Unknown definition in our data*2 |
| --- | --- | --- | --- | --- | --- |
| 1 | Myocardial infarction | I210-219, I220, I221, I228, I229, I252 | 52 | — | with Hospitalization and with ECG change and/or cardiac marker elevation |
|  | Congestive heart failure | I099, I110, I255, I420, I425-429, I430 (A188), I431 (E888), I431/E889, I432 (E639), I438 (E059), I500-509 | 44 | — | NYHA functional classification II to IV symptoms |
| | Peripheral vascular disease | I700-702, I708-716, I718, I731, I738, I739, I771, K551, K552, K558, K559 | 101 | — | with intermittent claudication or past bypass for chronic arterial insufficiency, history of gangrene or acute arterial insufficiency, or untreated thoracic or abdominal aneurysm ($\geq 6$ cm) |
|  | Dementia | B220, E756, F010-012, F019, F03, F051, F107, G10, G20, G238, G300, G301, G308-311, G318 | 28 | — | with chronic cognitive impairment |
|  | Cerebrovascular disease (including TIA) | G450, G451, G453, G454, G458, G459, H340, I600-611, I613-616, I618-621, I629-636, I638, I639, I64, I650-653, I660-663, I668-679, I690, I691, I693, I694 | 248 | — | with no or minor sequelae |
|  | Chronic pulmonary disease (including asthma) | I278, I279, J40-47, J60, J61, J628, J630-635, J64, J65, J660, J661, J670-679, J684, J701, J703 | 114 | — | — |
|  | Rheumatic disease (Lupus, polymyositis, mixed connective tissue disease, polymyalgia rheumatica, moderate to severe rheumatoid arthritis) | M0500, M0510, M0520, M0530, M0580-0588, M0590-0598, M0600-0608, M0610, M0620, M0630, M0640, M0680-0688, M0690-0698, M320, 321, 329-332, 339-341, M348-M351, M353 | 135 | — | — |
|  | Peptic ulcer disease | K250-K257, K259-267, K269, K270, K279, K284, | 60 | — | Any history of treatment for ulcer |
|  | (including ulcer bleeding) | K285, K287, K289 |  |  | disease |
| 2 | Hemiplegia/ paraplegia | G114, G801, G802, G810, G811, G819, G820-825, G830-834, G839 | 43 | — | — |
|  | Moderate or severe renal disease (including uremia) | I120, N032-034, N036, N037, N052-057, N183-185, N189, N19, N250 | 42 | — | Serum creatinine > 3.0mg/dL, dialysis or status post kidney transplant |
| 6 | AIDS (including HIV infection) | B24, B200, B202-206, B210-212, B220, B221, B238 | 17 | — | — |
| 1 | Mild liver disease (Chronic hepatitis or cirrhosis without portal hypertension) | B181, B182, B189, K700-703, K709, K713, K717, K730, 732, 738-741, K743-746, K760, K762-764, K768, K769 | 86 | — | — |
| 3 | Moderate and severe liver disease (Chronic hepatitis or cirrhosis with portal hypertension with/without variceal bleeding) | I850, I859, I864, K704, K711, K721, K729, K765, K766, K767 | 35 | — | — |
| 1 | Diabetes without chronic complication (Diabetes with medication) | E100, E101, E106, E109, E11, E119, E12, E13, E139, E14, E149, E831, E881, E888, E891 | 30 | including patients who were prescribed 487 oral hypoglycemic agent or 61 insulin preparation without insulin for intravenous injection (vial formulation) | — |
| 2 | Diabetes with chronic complication<br>—Retinopathy, neuropathy, or nephropathy | E102-105, E112-115, E132-135, E143-145, E888 | 41 | including patients who were prescribed drug for diabetes neuropathy (mexiletine hydrochloride) | — |
|  | — Hospitalized DKA or HHS | E100, E106, E116, E101, E111, E131, E140, E141, | 38 | including patients who prescribed |  |
|  |  | E110, E130 |  | insulin for intravenous injection<br>(vial formulation) |  |
| 2 | Tumor/ lymphoma/ leukemia | C000-004, C006, C008, C009, C01, C020-022,<br>C029-031, C039-041, C049-052, C059-062, C069-07, C080-081, C089-091, C100-104, C109-113,<br>C119, C12, C131,C132, C139, C140, C150-155,<br>C158-166, C169-172, C179-187, C189, C19, C20,<br>C210-211, C220-224, C227, C229, C23, C240,<br>C241, C248-254, C257-259, C261, C269-301,<br>C310-313, C319-323, C329, C33, C340-343,<br>C348-349, C37, C380-384, C400-403, C410-414,<br>C419, C430-447, C449-452, C459, C469-476,<br>C479-482, C490-496, C499, C500-506, C508-512,<br>C519, C52, C530, C531, C538-543, C549, C55,<br>C56, C570, C579, C58, C600-602, C609, C61,<br>C620, C621, C629-632, C637, C639, C64, C65,<br>C66, C670-677, C679, C681, C690-696, C700,<br>C701, C709, C710-721,C723-725, C729, C73,<br>C740, C741, C749-755, C760-765, C795, C809-814, C817, C819-821, C823, C824, C826, C827,<br>C829-831, C833, C835, C837, C838, C840, C841,<br>C844-848, C851, C852, C859-863, C865, C866,<br>C880, C882-884, C900-903, C910, C911, C913-925, C927-931, C933, C939, C940, C942-944,<br>C946, C947, C950, C951, C959, C960, C962,<br>C964-966, C968,<br>D000-002, D011-014, D019-023, D043-047,<br>D049-051, D059-061, D069-073, D090, D092, | 1738 | — | — |
|  |  | D099, D471, D475, D648, D728, D763, H350, L270 |  |  |  |
| 6 | Metastatic tumor | C770-775, C778, C779, C780-788, C790-799, C800, C809 | 158 | including patients with end-stage cancer and unknown primary cancer |  |

AIDS, Acquired immunodeficiency syndrome; DKA, Diabetic ketoacidosis; HHS, Hyperosmolar hyperglycemic syndrome; HIV, Human immunodeficiency virus; TIA, Transient ischemic attack; ICD-10, International Classification of Diseases, 10th revision; NYHA, New York Heart Association.
\*<sup>1</sup> Detail number of Japanese diagnostic code
\*<sup>2</sup> The definitions according to original Charlson Comorbidity Index [1]

## Statistical Analysis

All statistical analyses were conducted using the SAS-Enterprise Guide (SAS-EG 7.1) in SAS, version 9.4 (SAS Institute, Cary, NC, USA). A value of *p* < 0.05 was considered statistically significant.

## Results

***Table 2*** lists the clinical characteristics of the participants. The mean CCI of males, females, and both sexes combined were 0.67 ± 1.20, 0.68 ± 1.14, and 0.68 ± 1.17, respectively. The proportions of the CCI=0 group for males, females, and all subjects were 64.4% 61.5 %, and 63.1%, respectively. The proportions of the CCI ≥4 group were 3.69%, 3.00% and 3.38% in males, females, and in both sexes combined, respectively. In this study, males were younger and exhibited higher Body mass index, Systolic blood pressure, Diastolic blood pressure, Triglycerides, Aspartate aminotransferase, Alanine aminotransferase, γ-glutamyl transferase and Glycated hemoglobin values, and lower High-density lipoprotein cholesterol and Low-density lipoprotein cholesterol levels than females (all *p* < 0.0001). Furthermore, the CCI was higher in males than females (*p* < 0.0001). The most prevalent diseases of the 19 Charlson comorbidities were chronic pulmonary disease, followed by mild liver disease, and peptic ulcers. In females, the proportion of peptic ulcer disease was slightly higher than mild liver disease. Additionally, the proportions of peripheral vascular disease, dementia, chronic pulmonary disease, rheumatic disease, peptic ulcer disease, and tumor/lymphoma/leukemia were lower in males, whereas the proportion of other diseases was higher in males (all *p* < 0.0001).

**Table 2.**
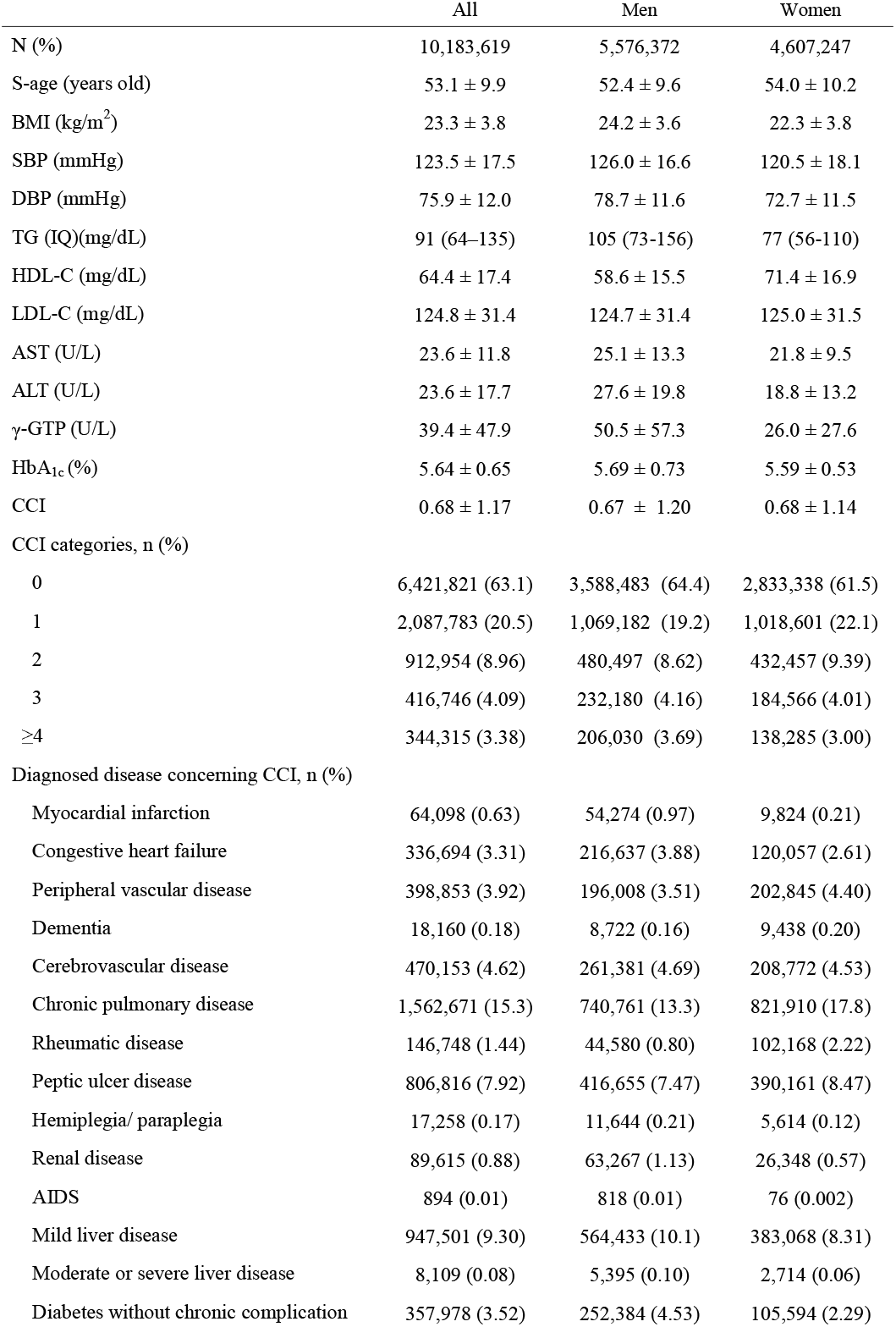
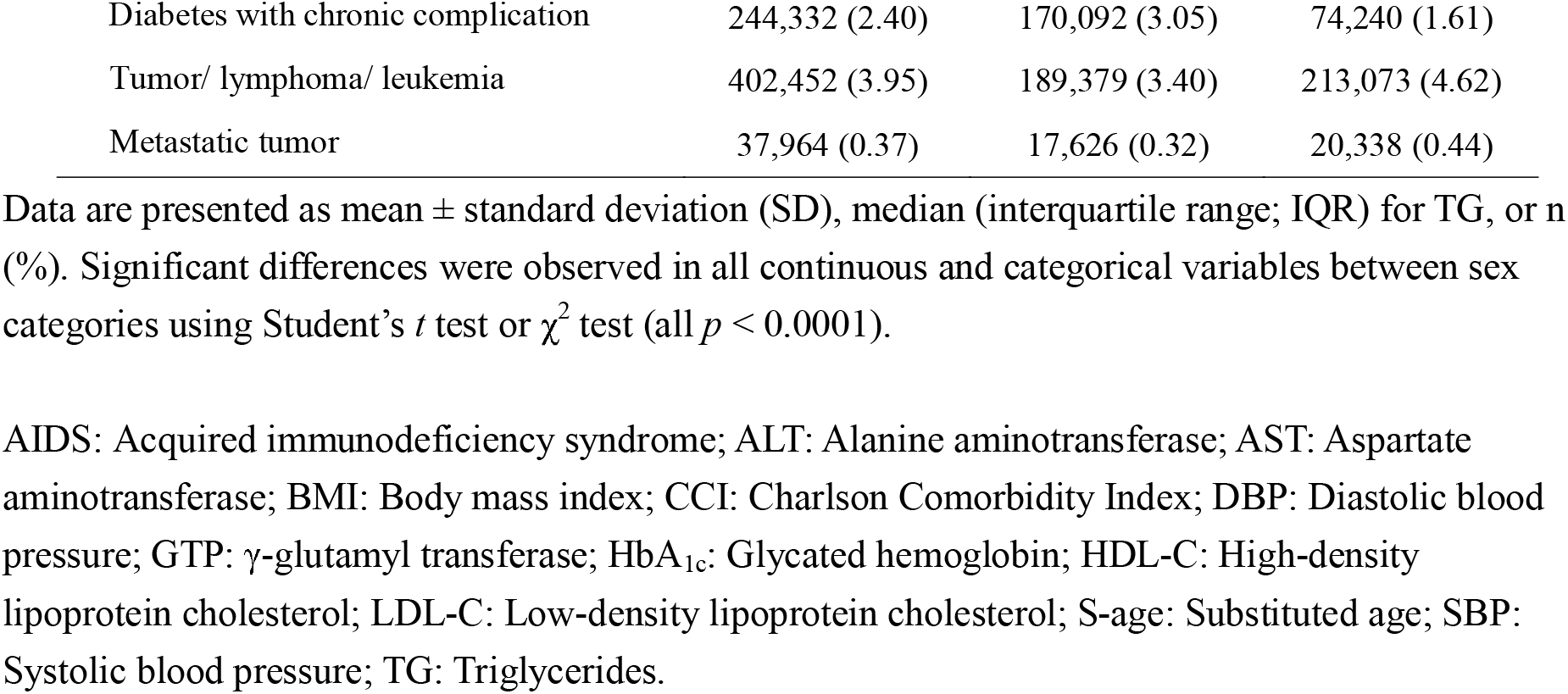
Clinical characteristics of participants.

The most prevalent comorbidity in patients with chronic pulmonary disease was asthmatic bronchitis (males: n = 461,854; 62.3%; females: n = 560,204; 68.2%). Similarly, the most prevalent comorbidity in patients with peptic ulcer disease was gastric ulcer (males: n = 331,600; 79.6%; females: n = 331,372; 84.9%). In patients with mild liver disease, the most prevalent comorbidity was a fatty liver in males (n = 239,453; 42.4%), whereas, in females, it was liver dysfunction (n = 165,066; 43.1%) followed by a fatty liver (n = 143,856; 37.6%).

## Discussion

We developed a new algorithm to define the CCI using diagnosed diseases and prescribed medications. Previous studies have also developed algorithms to define the CCI based on disease diagnosis codes using administrative claims databases [4,5]. However, as the original CCI included criteria that were not discernible from ICD-10 codes, these CCIs may have been excessively scored.

The CCI is the most widely used comorbidity index to estimate mortality rates in patients with multiple comorbidities and was originally used in longitudinal studies. Several studies have assessed the validity of the CCI as a predictor of mortality in patients with acute myocardial infarction [14], coronary artery disease [15], ischemic stroke [16], hemodialysis, peritoneal dialysis [17], postoperative esophageal cancer [18], and acute coronary syndrome [19], in cases of frailty in elderly surgical patients [20], and in the Medicare population aged 65 years or older [21]. Additionally, the CCI has been used in outcome studies to predict patient prognosis, with results suggesting that CCI is associated with the readmission of patients with heart failure [3] and postoperative complications (infections) from total hip arthroplasties [2].

A previous study using in-hospital discharge data suggested that five Charlson comorbidities (myocardial infarction, peripheral vascular disease, cerebrovascular disease, peptic ulcer disease, and diabetes without chronic complications) were not associated with in-hospital mortality and were excluded from the CCI criteria [22]. Moreover, various modified CCIs have been developed that have reweighed the index of comorbidities [23]. Therefore, further studies are needed to examine the association between our CCI and actual mortality in the latest database.

Our study had some limitations. First, several definitions of the CCI were unavailable in our database, including hospitalizations, detailed symptoms concerning ECG changes, NYHA functional classifications, serum creatinine levels (moderate: creatinine >3 mg/dL), dialysis, and post kidney transplant status (severe chronic kidney disease). Second, there was no available information concerning mortality in our database; therefore, it was impossible to confirm the association between our CCI and actual mortality. Additionally, as the mortality risk from comorbid diseases posed by an additional decade of age is equivalent to a one point increase in the CCI, Charlson et al. also developed an age-adjusted CCI for longitudinal studies [1]. Thus, further validation studies are needed to examine the association between age-adjusted CCIs and actual mortality in the NDB. However, our CCI algorithm-derivation process was more detailed than a previous method based on ICD-10 codes, whose validity was confirmed [5]. Notwithstanding these limitations, this study developed a detailed algorithm to define the CCI using diagnosed diseases and prescribed medication in the NDB-K7Ps. This CCI may be a useful tool for epidemiological studies using a real-world database and may provide novel insights into the comorbidity, pathophysiology, and underlying mechanisms for specific target diseases.

## Data Availability

Any inquiries regarding the availability of the data and the code to merge the datasets in this study should be directed to the corresponding author.

## Acknowledgments

We thank Tania Ellis, MPH, from Edanz (https://jp.edanz.com/ac) for editing a draft of this manuscript.

## Author Contributions

A.S. and K.N. contributed to the overall study design. A.S. and K.N. contributed to the interpretation of the initial results and a discussion of the literature. A.S. and K.N. prepared the data and software. A.S. prepared the first draft of the manuscript, and both authors read, edited, and approved the manuscript. All authors have read and agreed to the published version of the manuscript.

